# Daylight Saving Time, Acute Sleep Disruption, and Return-to-Baseline Patterns in All of Us Fitbit Data

**DOI:** 10.64898/2026.10.08.26365089

**Authors:** Shuai Yang, David A. Sbarra, Yann C. Klimentidis

## Abstract

Seasonal daylight saving time (DST) transitions abruptly alter the alignment between clock time, environmental light, and sleep. In preregistered analyses, we used longitudinal Fitbit data from the All of Us Research Program to compare sleep around 13 seasonal transitions in Colorado, New Mexico, and Utah with sleep on the same dates in Arizona, where clocks do not change. The sample included 1,522 participants and 6,658 participant-transition windows. Following the spring transition, sleep duration changed more from one night to the next in Mountain states than in Arizona (14.6 minutes, 95% CI [7.2, 22.0]), even though sleep duration itself on day +1 showed little evidence of a DST-related reduction (-8.4 minutes, 95% CI [-17.3, 0.6]). Sleep timing shifted later after the spring transition and about 28 minutes earlier after the fall transition. Among Mountain-state observations with a large post-transition departure from usual sleep duration, the median return to the usual range was two nights. Nearly the same two-night pattern appeared in Arizona after similarly large sleep departures, indicating that this return pattern was not specific to DST. Greater baseline sleep-duration variability predicted a greater likelihood of remaining outside the usual range at days +7 and +14. Seasonal clock changes produced acute sleep disruption, while the Arizona comparison shows that a rapid return to baseline should not be interpreted as the duration of recovery from DST itself.

**Statement of Significance:** Seasonal daylight saving time changes are often studied with before-and-after comparisons, which can mix clock-change effects with ordinary variation across the week. Using Arizona as a calendar-matched no-clock-change comparison, we found that the Mountain-versus-Arizona difference in sleep duration on spring day +1 was similar to that on day -1, while the change from the preceding observed night was larger in Mountain states. In a separate Mountain-state analysis, large post-transition departures usually returned to the pre-transition ±30-minute window within two nights. The same return pattern appeared on the same spring and fall dates in Arizona as well. This distinction shows why acute exposure effects and threshold-defined return trajectories should be evaluated separately in wearable studies.

## INTRODUCTION

Seasonal daylight saving time (DST) advances civil time by one hour in spring and returns to standard time in fall. Solar time and the light-dark cycle do not move with the clock, so the transition abruptly changes the relation between environmental light, social schedules, and internal circadian timing. This mismatch can alter sleep opportunity and sleep timing, especially after the spring advance.^1,2^ Debate continues over whether seasonal clock changes should be eliminated and, if so, whether permanent standard or daylight saving time should be adopted. The American Academy of Sleep Medicine and the Sleep Research Society favor permanent standard time, based largely on circadian alignment and concerns about the acute effects of clock changes on sleep and health.^3,4^ Evidence for downstream health effects, however, remains mixed across outcomes and study designs, as highlighted in a recent systematic review of 157 studies.^5^ Sleep provides a proximal behavioral measure of the acute clock transition and can be followed night by night after the change.

Wearable and device-based studies generally report their largest sleep changes after the spring transition, although estimates vary. Lahti et al.^6^ observed shorter sleep after the spring change, and Owen et al.^7^ reported an acute reduction of nearly 30 minutes in estimated sleep period in the Project Baseline Health Study. Heacock et al.^8^ detected changes in sleep timing and duration around U.S. clock transitions in more than 24,000 wearable users. Large wearable cohorts also show strong weekday and seasonal structure in sleep, including longer weekend sleep.^9,10^ A systematic review of 27 daylight saving time sleep studies concluded that spring transitions tend to adversely affect sleep duration or quality while emphasizing methodological inconsistency and external calendar influences.^2^ Many studies rely on before-and-after comparisons within the same population, leaving ordinary day-of-week and seasonal changes as plausible contributors. A contemporaneous no-DST comparison can separate those calendar effects from the local clock change.

Sleep duration on a single night and change between nights capture different features of sleep. For example, 6.5 hours on one night followed by 7.5 hours on the next gives a two-night mean of 7 hours, while the night-to-night change is 60 minutes. Sleep regularity has emerged as a dimension of sleep health that can carry information beyond mean duration.^11–13^ This motivates examining both nightly duration and the absolute change in nightly sleep duration from one observed night to the next. Return-to-baseline measures require a separate concern. Selecting a large departure from a recent baseline and then waiting for subsequent values to approach the same baseline creates susceptibility to regression to the mean.^14^ A no-DST specificity analysis can show whether the same return pattern appears after large departures that occur without a local clock change.

The All of Us Research Program provides multi-year Fitbit observations across repeated seasonal transitions.^15,16^ We paired Arizona, where clocks do not change, with the neighboring DST-observing Mountain states of Colorado, New Mexico, and Utah. Jeong et al.^17^ recently used this regional contrast to study physical activity and resting heart rate and found prominent shifts in activity timing with little net change in total daily steps. Their analysis did not evaluate nightly sleep. The present study extends this regional natural experiment to sleep across 13 separate spring and fall clock-change transitions and examines both acute nightly changes and individual return to a recent sleep-duration baseline.

We preregistered three aims on the Open Science Framework (https://osf.io/52ucq/registrations). Aim 1 tested whether sleep changed around the clock transition in DST-observing Mountain states relative to calendar-matched changes in Arizona. The primary outcomes were nightly sleep duration and the absolute change in sleep duration from one observed night to the next; sleep efficiency and sleep timing were secondary outcomes. Aim 2 described how long sleep duration took to return to the pre-transition ±30-minute window among Mountain participant-transitions with a >30-minute departure on days +1 to +3 after a DST transition. Aim 3 tested whether short baseline sleep or greater baseline sleep-duration variability predicted remaining outside that window at days +7 and +14. After completing the primary analyses, we applied the Aim 2 selection-and-return algorithm to Arizona as a post hoc specificity check. Its purpose was to determine whether the observed return curve required a local clock change or could also arise after similarly large departures on matched dates without a clock change. Additional post hoc checks examined weekday patterning and departure direction.

## METHODS

### Participants and data source

We conducted a secondary analysis of Curated Data Repository version 8 (CDR v8) Controlled Tier data from the National Institutes of Health All of Us Research Program. Participant data were restricted to records available by October 1, 2023. All row-level extraction, sleep processing, and modeling occurred inside the secure All of Us Researcher Workbench; only aggregate results meeting dissemination requirements were exported. The analytic cohort included adults with Fitbit sleep data around at least one eligible transition and a participant-level state-of-residence field placing them in Arizona, Colorado, New Mexico, or Utah.

State of residence came from the participant-level cb_search_person.state_of_residence field. We used the value available in the Curated Data Repository as a fixed classification across observed transition windows because transition-specific residence dates were unavailable. Arizona formed the calendar-matched no-DST comparison, and Colorado, New Mexico, and Utah formed the DST-observing Mountain group. Age at each transition was calculated from transition year and year of birth. A participant-transition was one person observed around one spring or fall clock change. The calendar contained 13 eligible transitions from spring 2017 through spring 2023, and participants could contribute multiple windows.

Primary eligibility required at least seven valid nights during days -14 to -1 and at least seven valid nights during days +1 to +14. Aim 1 used nightly observations nested within participant-transitions. Aims 2 and 3 used one record per eligible participant-transition after applying the qualifying-departure rule described below. Repeated windows from the same participant were retained and accounted for in inference.

### DST windows and nightly sleep construction

Spring daylight saving time was defined as the second Sunday in March and fall daylight saving time as the first Sunday in November. Event day 0 was the local transition date. Nightly records from day -21 through day +21 were assembled for processing and diagnostics; primary models used days -14 to -1 and +1 to +14. Day 0 was excluded from the primary event-study sample because the clock changes during the usual sleep period and complicates nightly date assignment. The same calendar dates were used in Arizona.

We reconstructed nightly sleep from Fitbit sleep_level records with the published typical sleep period (TSP) algorithm implemented in the typical.sleep R package.^18^ The algorithm assigns fragmented stage-level records to a participant’s habitual primary sleep episode. Source data began 120 days before the first eligible transition to provide look-back history and extended through 21 days after the last transition. The 120-day interval was a processing window and carried no eligibility requirement. A valid night contained 2 to 15 hours of sleep and sleep efficiency from 50% to 100%. A prespecified sensitivity used Fitbit’s default isMainSleep summary. The exact processing sequence and field mappings are provided in the Supplementary Methods.

### Measures and analysis

Baseline measures were calculated separately within each participant-transition from valid nights on days -14 through -1, the 14-day pre-transition window. Baseline sleep duration was the mean of all valid nights available in that window; primary eligibility required at least seven valid pre-transition nights. The standard deviation of those nightly durations quantified baseline variability, and short baseline sleep was defined as a mean below 6 hours. Mean sleep efficiency was calculated over the same window. For clock-time measures, the TSP onset, midpoint, and wake fields were converted to minutes after midnight using circular calculations described in the Supplementary Methods.

Aim 1 used two primary outcomes across every valid night in the event-study window (days -14 to -1 and +1 to +14). Nightly sleep duration was measured as total sleep time in minutes. Night-to-night change was the absolute difference between the current observed night’s sleep duration and the preceding observed night’s duration after chronological ordering within each participant-transition. For example, a 6.5-hour night followed by a 7.5-hour night yields a 60-minute night-to-night change, regardless of direction. Day +1 was the focal post-transition estimate, although the outcome was calculated throughout the event-study window. Day 0 was excluded as its own event-study observation. Because night-to-night change was calculated on the chronologically ordered nightly series before the event-study filter was applied, an observed day 0 night could serve as the preceding night for the day +1 outcome. The preregistered measure used the preceding observed night and could span a missing calendar night. Sleep efficiency and sleep timing were secondary outcomes. Absolute deviation from the participant-transition baseline was examined as an unconditioned supporting outcome.

For each season, Aim 1 used an event-study difference-in-differences model with participant-transition and exact-date fixed effects; standard errors were clustered by participant. Event day -1, the Saturday immediately before the Sunday clock-change date, was the reference. Event day 0 was Sunday, and day +1 was Monday. Each coefficient compares the Mountain-versus-Arizona difference on a given event day with the corresponding difference on Saturday, day -1. Spring and fall were estimated separately, and days -14 through -2 were tested jointly for differential pre-transition trends.^19,20^ Because the focal day +1 night-to-night estimate compares a Monday observation with its preceding observed night, we added a post hoc sensitivity restricted to consecutive Sunday-to-Monday observations. It compared the Mountain-versus-Arizona difference on post-transition Monday with the corresponding differences for the two ordinary pre-transition Sunday-to-Monday pairs, using Mondays at days -13 and -6 as the comparison points. Both nights in each pair had to be valid and exactly one calendar day apart. Full formulas appear in the Supplementary Methods.

Aim 2 described return to baseline among Mountain participant-transitions that met the 7-pre/7-post rule. A window qualified when at least one valid night on days +1 to +3 differed from its mean pre-transition sleep duration by more than 30 minutes in either direction. The 30-minute cutoff was the preregistered operational definition of a large departure. The first qualifying departure was the earliest such night. Follow-up began after that night, and return was the first subsequent valid night through day +14 with sleep duration within ±30 minutes of the pre-transition mean. Windows without an observed return were censored at day +14. Cox proportional hazards models related the absolute magnitude of the first qualifying departure to time to return and accounted for repeated participant-transitions within participants. After completing the preregistered Mountain-state analysis, we applied the identical selection-and-return algorithm to calendar-matched Arizona windows as a post hoc specificity check. This comparison asked whether the observed return curve required a local clock change. A similar curve in Arizona would indicate that ordinary within-person variability and regression to the mean contributed substantially to the threshold-defined return pattern. If these large post-transition departures reflected a DST-specific disruption, we expected their subsequent return pattern to differ from similarly large departures observed in Arizona on the same calendar dates. Additional post hoc checks examined departure direction and recalculated each day +1 to +3 departure using the same weekday from the two pre-transition weeks as the reference.

Aim 3 examined whether pre-transition sleep characteristics predicted remaining outside the baseline ±30-minute window among qualifying Mountain participant-transitions. Separate logistic regression models evaluated this outcome at transition day +7 and day +14. Primary predictors were baseline short sleep (<6 hours) and baseline sleep-duration variability; variability and age were standardized. Models also included state and transition type. Participant-clustered robust Wald standard errors accounted for repeated transitions.

Full model specifications and post hoc sensitivity definitions are provided in the Supplementary Methods; the preregistration crosswalk appears in Supplementary Table S1. Event-study models were fitted with the *fixest* R package,^21^ survival models with the *survival* R package,^22^ and sleep preprocessing with *typical.sleep*.^18^ Analyses were conducted in R 4.5.0 with two-sided tests and 95% confidence intervals. Post hoc analyses are identified as such throughout.

## RESULTS

### Analytic sample

The TSP pipeline identified 1,926 Fitbit sleep contributors with 294,691 nightly records across the stacked daylight saving time windows, forming 9,363 participant-transitions. The primary completeness rule retained 1,522 participants and 6,658 participant-transitions: 697 participants in Arizona and 825 in the Mountain states. Within the Mountain group, 3,023 windows were eligible for the return analysis before the acute-departure criterion, and 2,689 windows from 799 participants qualified (Figure 1). Mean baseline sleep duration was 7.12 hours and mean baseline sleep efficiency was 90.4% (Table 1).

**Figure 1.**
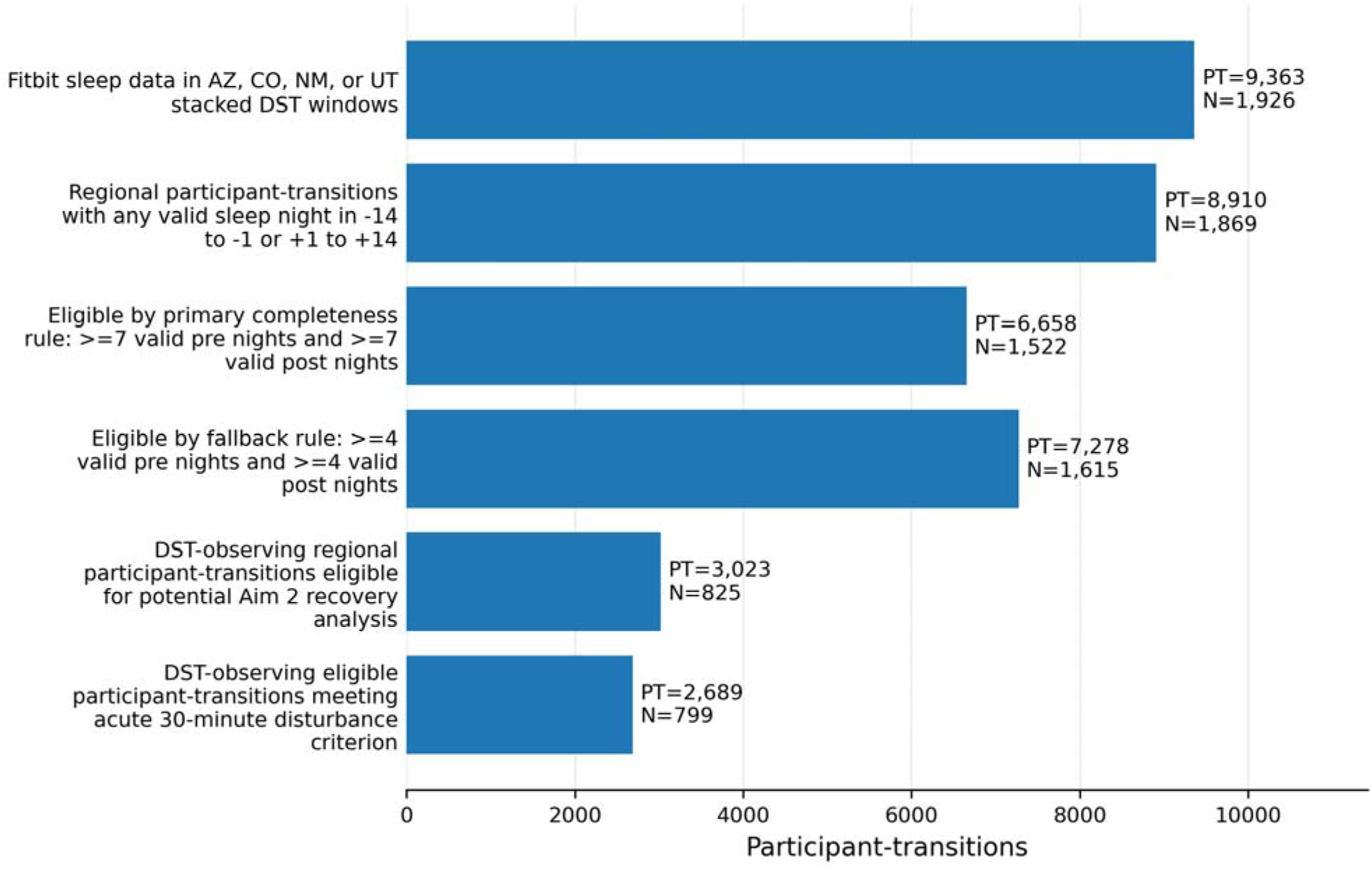
Analytic sample flow. A participant-transition is one participant observed around one spring or fall clock change; a participant can contribute multiple transition windows. N denotes unique participants and PT denotes participant-transitions. Alt text: Flow diagram showing construction of the analytic sample from Fitbit sleep contributors through participant-transition eligibility and the final Aim 2 qualifying cohort. Counts are shown for unique participants and participant-transition windows.

**Table 1.** Characteristics of participants and eligible DST transition windows.

| Characteristic | Level | Overall | Arizona no-DST | Mountain DST |
| --- | --- | --- | --- | --- |
| Participants |  | 1,522 | 697 | 825 |
| Participant-transitions |  | 6,658 | 3,635 | 3,023 |
| Age at transition, years |  | 56.0 (15.2) | 57.4 (14.9) | 54.3 (15.3) |
| Baseline sleep duration, hours |  | 7.12 (0.97) | 7.09 (1.00) | 7.16 (0.92) |
| Baseline sleep efficiency, % |  | 90.4 (3.8) | 90.4 (3.9) | 90.4 (3.6) |
| Baseline sleep-duration SD, hours |  | 1.18 (0.50) | 1.20 (0.50) | 1.16 (0.50) |
| Baseline short sleep <6 h |  | 780 (11.7%) | 468 (12.9%) | 312 (10.3%) |
| Transition type | Fall | 3,046 (45.7%) | 1,683 (46.3%) | 1,363 (45.1%) |
| Transition type | Spring | 3,612 (54.3%) | 1,952 (53.7%) | 1,660 (54.9%) |
| State | AZ | 3,635 (54.6%) | 3,635 (100.0%) |  |
| State | CO | 1,980 (29.7%) |  | 1,980 (65.5%) |
| State | NM | 580 (8.7%) |  | 580 (19.2%) |
| State | UT | 463 (7.0%) |  | 463 (15.3%) |
| Gender | Female | 4,547 (68.3%) | 2,583 (71.1%) | 1,964 (65.0%) |
| Gender | Male | 1,935 (29.1%) | 979 (26.9%) | 956 (31.6%) |
| Gender | Another identity, multiple, or not reported | 176 (2.6%) | 73 (2.0%) | 103 (3.4%) |
| Race | White | 5,588 (83.9%) | 3,045 (83.8%) | 2,543 (84.1%) |
| Race | Black or African American | 116 (1.7%) | 84 (2.3%) | 32 (1.1%) |
| Race | Other, multiple, or not reported | 954 (14.3%) | 506 (13.9%) | 448 (14.8%) |
| Ethnicity | Hispanic or Latino | 637 (9.6%) | 308 (8.5%) | 329 (10.9%) |
| Ethnicity | Not Hispanic or Latino | 5,879 (88.3%) | 3,254 (89.5%) | 2,625 (86.8%) |
| Ethnicity | Other or not reported | 142 (2.1%) | 73 (2.0%) | 69 (2.3%) |

Note. One participant can contribute several transition windows. Except for the unique-participant row, values and percentages summarize participant-transition windows. Age was updated for each transition year. Sparse demographic categories were combined before export to meet All of Us small-cell dissemination requirements; analytic models used the original variables. SD = standard deviation.

### Aim 1: Acute changes in sleep around daylight saving time

The first primary outcome was sleep duration on the event night itself. On spring day +1, the Mountain-versus-Arizona difference in sleep duration was similar to that on day -1 (estimate = -8.4 minutes, 95% CI [-17.3, 0.6], p = .066). The corresponding fall estimate was -3.4 minutes (95% CI [-12.7, 5.9], p = .473). The Mountain-versus-Arizona difference in sleep efficiency on day +1 was likewise similar to that on day -1 (Table 2). Overall, there was little evidence that the spring or fall clock transition altered the amount of sleep obtained on day +1 relative to the Arizona comparison condition.

**Table 2.** Day +1 event-study estimates for primary and secondary sleep outcomes.

| Season | Outcome | Estimate (95% CI) | p | N | PT |
| --- | --- | --- | --- | --- | --- |
| Spring | Sleep duration, min | -8.4 (-17.3 to 0.6) | .066 | 1,391 | 3,612 |
| Spring | Sleep efficiency, percentage points | 0.15 (-0.40 to 0.70) | .601 | 1,391 | 3,612 |
| Spring | Absolute change from preceding observed night, min | 14.6 (7.2 to 22.0) | <.001 | 1,391 | 3,612 |
| Spring | Sleep onset change, min | 17.3 (9.3 to 25.2) | <.001 | 1,391 | 3,612 |
| Spring | Sleep midpoint change, min | 12.5 (5.7 to 19.4) | <.001 | 1,391 | 3,612 |
| Spring | Wake time change, min | 8.0 (-1.0 to 16.9) | .081 | 1,391 | 3,612 |
| Fall | Sleep duration, min | -3.4 (-12.7 to 5.9) | .473 | 1,264 | 3,046 |
| Fall | Sleep efficiency, percentage points | -0.00 (-0.58 to 0.58) | .998 | 1,264 | 3,046 |
| Fall | Absolute change from preceding observed night, min | 2.5 (-5.6 to 10.5) | .551 | 1,264 | 3,046 |
| Fall | Sleep onset change, min | -26.7 (-35.8 to -17.7) | <.001 | 1,264 | 3,046 |
| Fall | Sleep midpoint change, min | -28.1 (-35.5 to -20.7) | <.001 | 1,264 | 3,046 |
| Fall | Wake time change, min | -27.9 (-37.0 to -18.8) | <.001 | 1,264 | 3,046 |

Note. Estimates compare Mountain daylight saving time states with Arizona at day +1 relative to day -1. Participants are counted separately within season. Many participants contributed both spring and fall windows, so the 1,391 spring participants and 1,264 fall participants overlap; the participant-transition counts sum to the 6,658-window primary sample. Timing analyses required at least four valid pre-transition timing observations. CI = confidence interval; N = participants; PT = participant-transitions.

The second primary outcome was the absolute change in sleep duration from the preceding observed night to the event night. For spring day +1, this night-to-night change was 14.6 minutes larger in Mountain states than in Arizona relative to day -1 (95% CI [7.2, 22.0], p < .001; Figure 2). The fall estimate was 2.5 minutes (95% CI [-5.6, 10.5], p = .551). The spring pre-transition coefficients showed no joint regional divergence for this outcome (p = .654; Supplementary Table S2). That is, the larger night-to-night change following the spring transition remained when the analysis was restricted to consecutive Sunday-to-Monday nights and compared with ordinary Sunday-to-Monday changes before the transition (Supplementary Table S3). Overall, the spring transition was associated with a larger-than-usual change in sleep duration from one night to the next, whereas no corresponding effect was evident following the fall transition.

**Figure 2.**
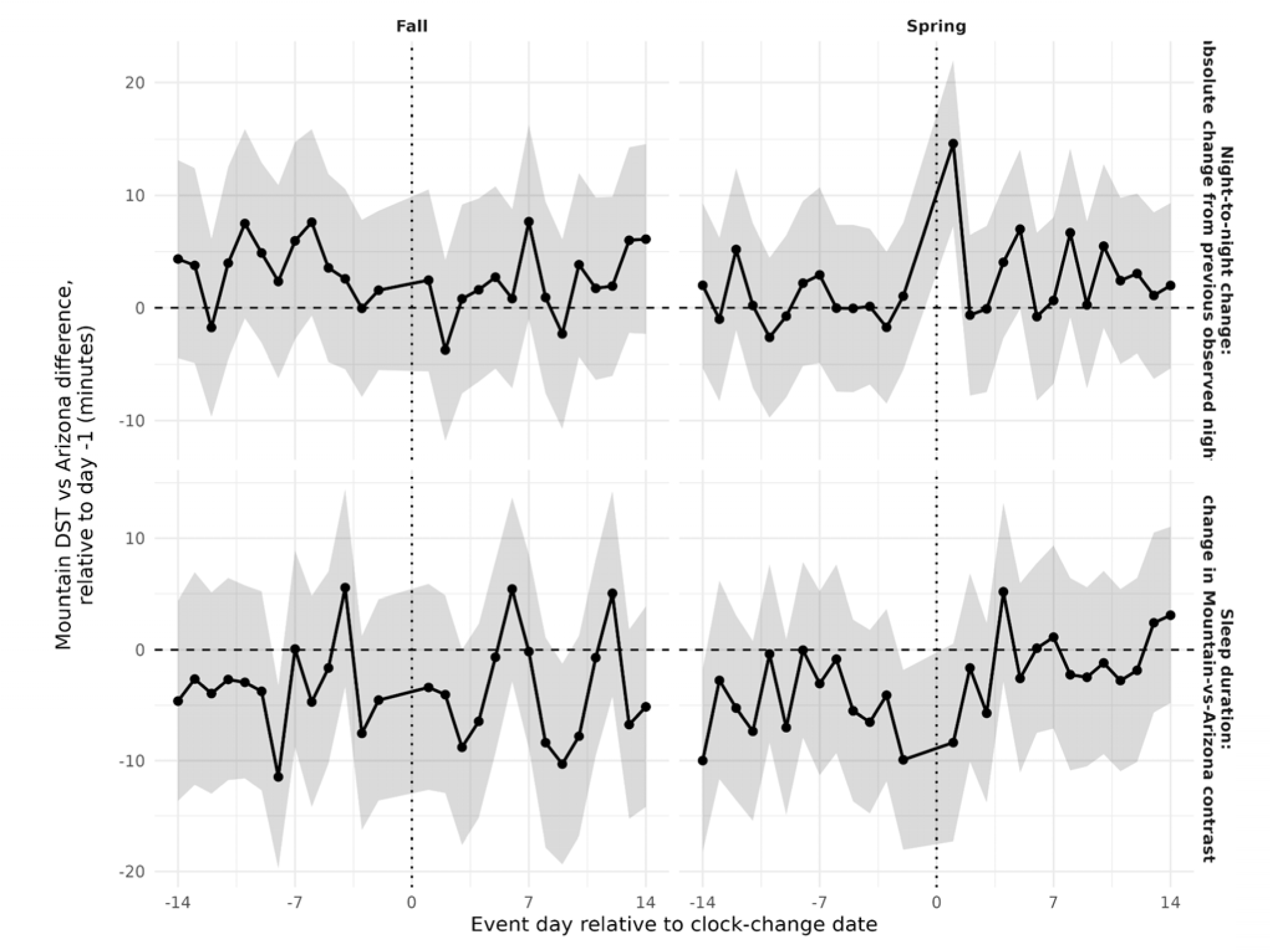
Event-study estimates for the two primary Aim 1 outcomes. The bottom row shows sleep duration on the event night itself. The top row shows the absolute difference between that night’s sleep duration and the preceding observed night’s duration. For day +1, an observed day 0 night could provide the preceding-night value even though day 0 was not included as its own event-study point. For example, 20 minutes means the two observed nights differed by 20 minutes, regardless of which night was longer. Each point compares the Mountain-versus-Arizona difference on that event day with the corresponding difference on Saturday, event day -1. Shaded bands are 95% confidence intervals. The vertical dotted line marks the Sunday clock-change date, event day 0. Alt text: Four-panel event-study plot with fall and spring columns. The bottom row shows sleep duration on each event night, and the top row shows the absolute change from the preceding observed night. The spring day +1 night-to-night estimate is clearly positive, while the fall estimate is near zero.

### Aim 2: Return to baseline after a large post-transition departure

Among the 3,023 eligible Mountain transition windows, 2,689 (88.9%) met the >30-minute qualifying-departure criterion. The first qualifying departure occurred on day +1 in 93.8% of spring windows and 94.5% of fall windows. Median time from the first qualifying departure to the first subsequent return within the baseline ±30-minute window was two nights in both seasons. By transition day +7, 17.1% of spring windows and 16.2% of fall windows remained outside the window (Table 3; Figure 3). Alternative qualifying definitions produced similar return summaries (Supplementary Table S4).

**Figure 3.**
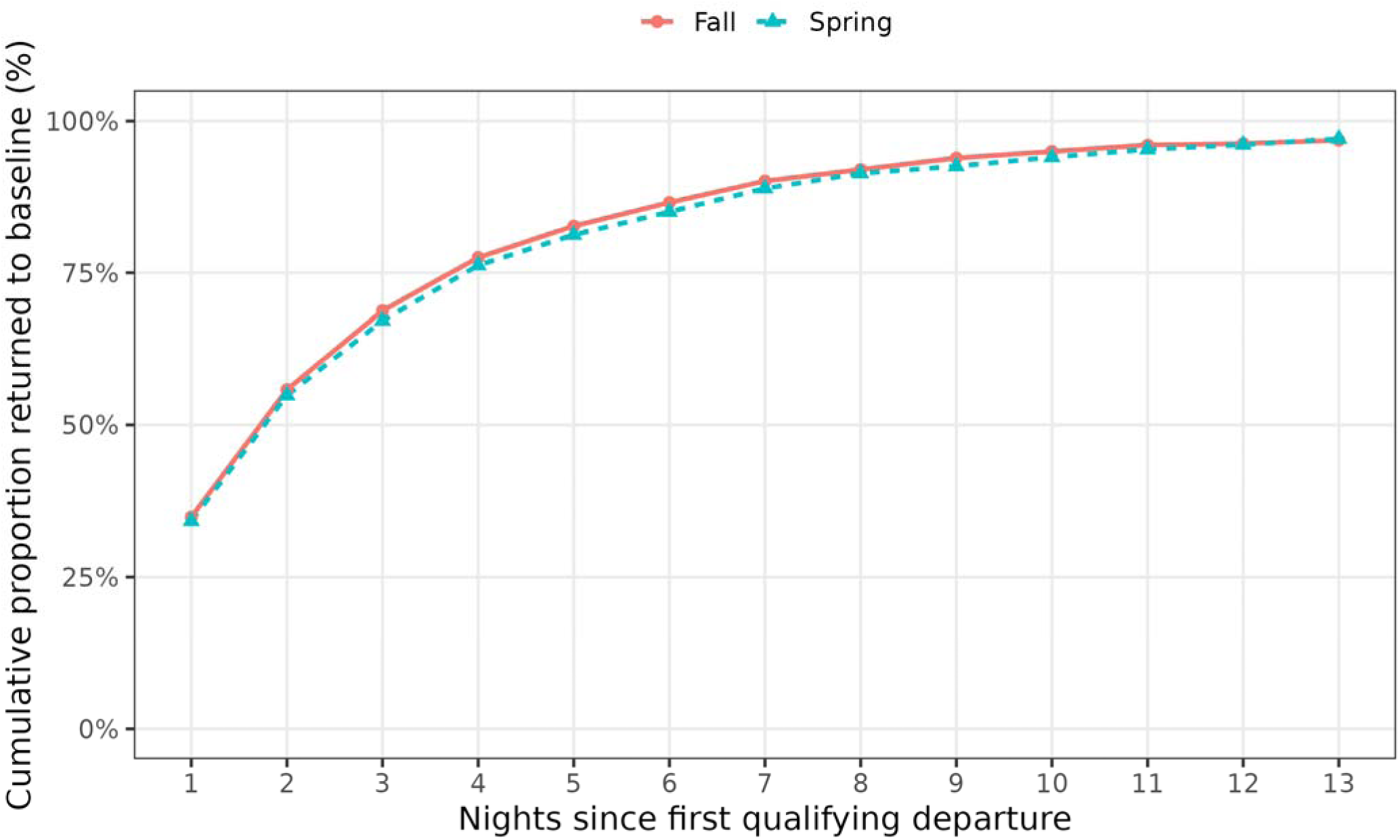
Cumulative return to baseline sleep duration after the first qualifying post-transition departure in DST-observing Mountain states. The x-axis is nights since the first valid day +1 to +3 with an absolute sleep-duration departure >30 minutes from the participant-transition baseline; return requires a subsequent valid night within 30 minutes of baseline. Alt text: Cumulative return curves for qualifying Mountain-state windows in spring and fall. Both curves rise rapidly, with a median return of two nights and about 16% to 17% of windows still outside the baseline range at day +7.

**Table 3.** Return to baseline sleep and predictors of delayed return.

| Analysis | Group/outcome | Measure | Estimate |
| --- | --- | --- | --- |
| Return summary | Fall | Median nights from first qualifying departure to recovery | 2 nights |
| Return summary | Spring | Median nights from first qualifying departure to recovery | 2 nights |
| Return summary | Fall | Unrecovered by transition day +7 | 16.2% |
| Return summary | Spring | Unrecovered by transition day +7 | 17.1% |
| Cox return model | All | Baseline short sleep <6 h | HR 0.80 (0.67 to 0.97); p = .020 |
| Cox return model | All | First qualifying departure magnitude, per SD | HR 0.84 (0.80 to 0.87); p = <.001 |
| Delayed-return logistic | Day +7 | Baseline short sleep <6 h | OR 1.56 (1.13 to 2.15); p = .007 |
| Delayed-return logistic | Day +7 | Baseline sleep-duration SD, per SD | OR 1.77 (1.60 to 1.95); p = <.001 |
| Delayed-return logistic | Day +14 | Baseline short sleep <6 h | OR 1.78 (0.99 to 3.18); p = .053 |
| Delayed-return logistic | Day +14 | Baseline sleep-duration SD, per SD | OR 1.94 (1.66 to 2.26); p = <.001 |

Note. Time to return begins at the first qualifying >30-minute post-transition departure and requires a subsequent valid night within 30 minutes of the participant-transition baseline. HRs below 1 indicate slower subsequent return; ORs above 1 indicate greater odds of remaining unrecovered at the stated transition-day checkpoint. Logistic-model intervals and p values use participant-clustered robust Wald inference. HR = hazard ratio; OR = odds ratio; SD = standard deviation.

Larger first departures were followed by slower return. Each one-standard-deviation increase in absolute first-departure magnitude was associated with a 16% lower return hazard (hazard ratio [HR] = 0.84, 95% CI [0.80, 0.87], p < .001). Short baseline sleep was also associated with slower return (HR = 0.80, 95% CI [0.67, 0.97], p = .020).

The post hoc specificity checks asked whether the same threshold-defined pattern would appear on matched Arizona dates without a local clock change. Qualifying departures were similarly common in Arizona: 88.4% of spring windows and 90.3% of fall windows qualified, compared with 88.9% and 89.0% in Mountain states. First qualifying departures were nearly evenly split between shorter and longer sleep in both regions (Supplementary Table S5). The median return was two nights in Arizona as well, and the adjusted Mountain-versus-Arizona return hazard ratio was 1.03 (95% CI [0.98, 1.09], p = .247; Supplementary Table S6; Figure S1). The unconditioned absolute-deviation event study likewise showed no clear post-transition excess in Mountain states (Figure S2). Replacing the overall pre-transition mean with a same-weekday reference left qualifying rates above 91% in every region-season stratum (Supplementary Table S7). These checks show that the two-night return curve was not specific to the local daylight saving time transition. Contrary to the expectation that DST-related circadian disruption might produce a more prolonged recovery than ordinary fluctuations in sleep, a similar return pattern was observed in Arizona. Thus, the two-night estimate should not be interpreted as the time required to recover from DST-related sleep or circadian disruption; it describes a return pattern that also occurs after large sleep departures without a local clock change.

### Aim 3: Baseline predictors of remaining outside the baseline window

At transition day +7, short baseline sleep was associated with higher odds of remaining outside the baseline ±30-minute window (odds ratio [OR] = 1.56, 95% CI [1.13, 2.15], p = .007). Each one-standard-deviation increase in baseline sleep-duration variability was associated with higher odds of remaining outside the window (OR = 1.77, 95% CI [1.60, 1.95], p < .001). At day +14, baseline variability remained associated with the outcome (OR = 1.94, 95% CI [1.66, 2.26], p < .001); the short-sleep estimate was OR = 1.78 (95% CI [0.99, 3.18], p = .053; Table 3; Figure 4). Full coefficients are reported in Supplementary Table S8.

**Figure 4.**
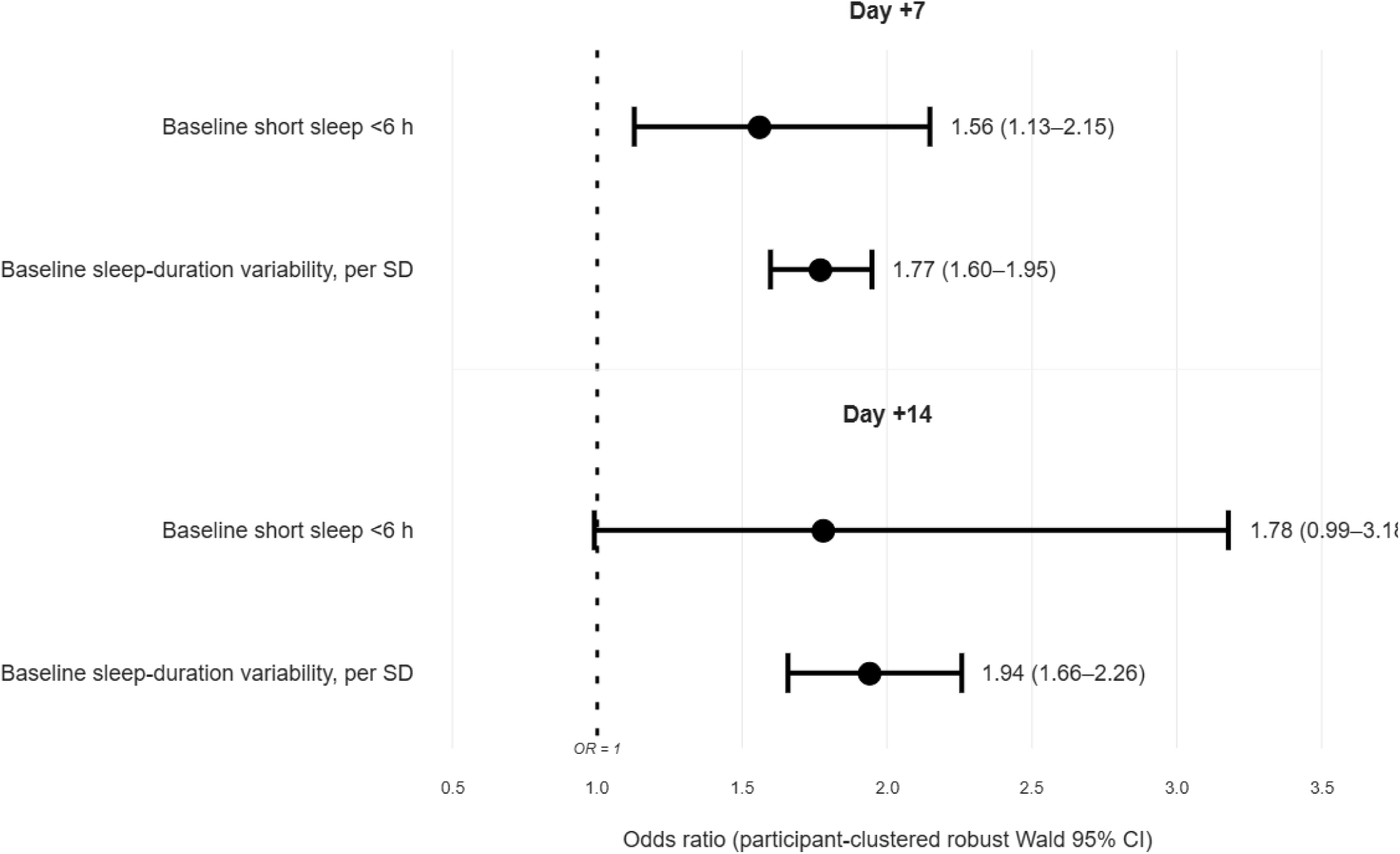
Baseline predictors of remaining outside the pre-transition sleep-duration ±30-minute window at transition day +7 or +14. Points are odds ratios from the parsimonious logistic models and whiskers are participant-clustered robust Wald 95% confidence intervals. The dotted vertical line marks an odds ratio of 1. Alt text: Forest plot of odds ratios for short baseline sleep and baseline sleep-duration variability at days +7 and +14. Greater baseline variability is associated with higher odds of remaining outside the baseline window at both checkpoints; the day +14 short-sleep interval includes the null.

### Secondary Aim 1 outcome: sleep timing

Sleep timing also shifted around the clock changes. On spring day +1, sleep onset was 17.3 minutes later in Mountain states relative to Arizona (95% CI [9.3, 25.2], p < .001), and midpoint was 12.5 minutes later (95% CI [5.7, 19.4], p < .001). Wake time was 8.0 minutes later (95% CI [-1.0, 16.9], p = .081). In fall, onset, midpoint, and wake time were 26.7, 28.1, and 27.9 minutes earlier, respectively (all p < .001; Table 2). Figure 5 shows the full timing trajectories.

**Figure 5.**
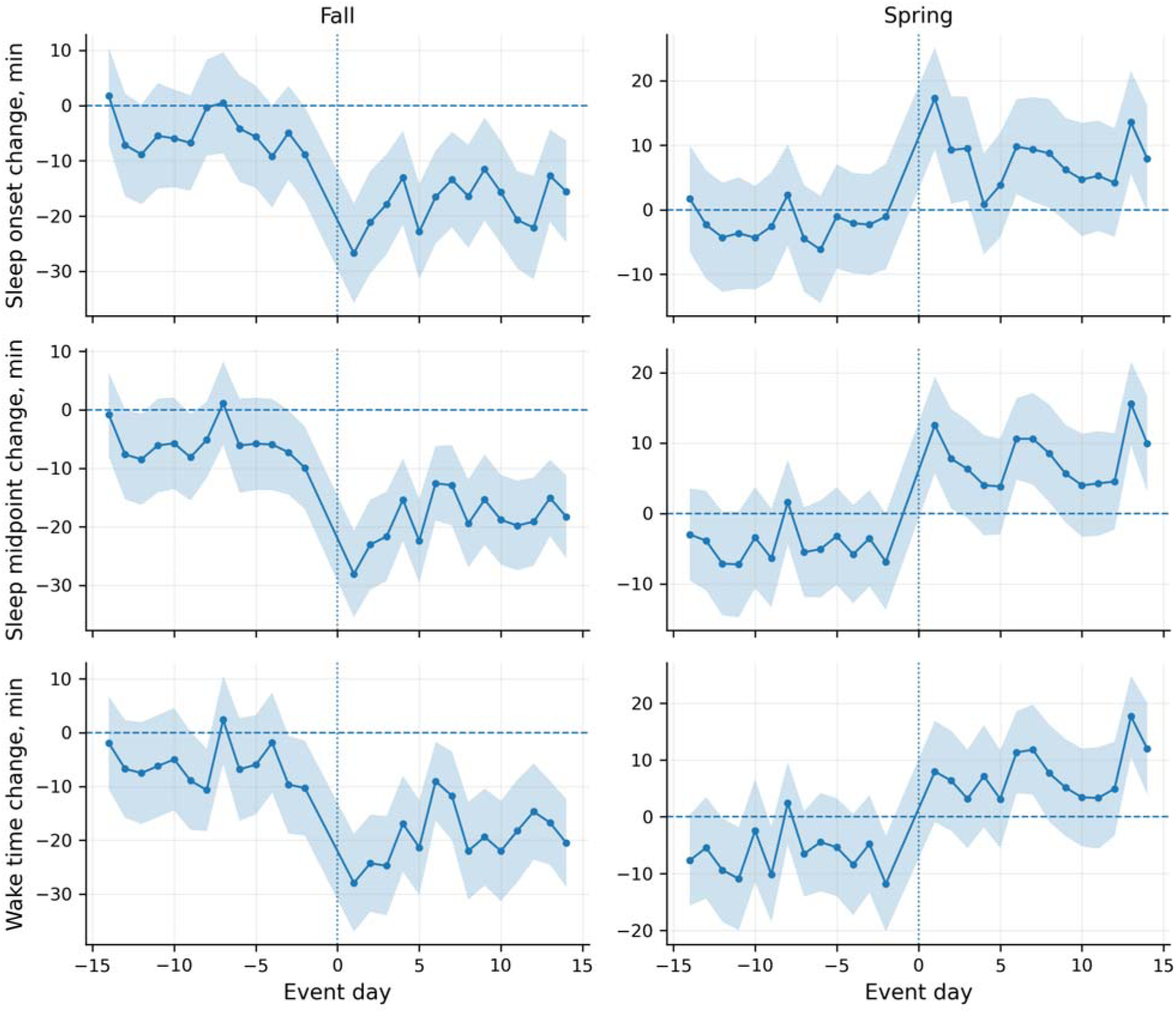
Secondary event-study estimates for sleep onset, midpoint, and wake time. Positive values indicate later clock time and negative values indicate earlier clock time relative to the participant-transition baseline. Alt text: Event-study plots for sleep onset, midpoint, and wake time. Spring estimates shift later after the transition, while fall estimates shift earlier by roughly half an hour on day +1.

### Additional sensitivity analyses

Sensitivity analyses examined whether the primary findings were robust to alternative sleep-processing decisions and the exclusion of the COVID-19 pandemic period. First, using Fitbit’s default isMainSleep pipeline rather than the typical sleep period algorithm increased the analytic sample to 2,021 participants and 8,666 eligible transition windows. Under this alternative pipeline, the spring day +1 night-to-night estimate was smaller than in the primary analysis (7.0 minutes, 95% CI [-0.3, 14.2], p = .060, compared with 14.6 minutes; Supplementary Table S9). Second, after excluding the 2020 and 2021 transitions, the spring estimate remained positive at 10.3 minutes (95% CI [1.7, 18.9], p = .018), and the day +7 return proportions remained close to the primary estimates (Supplementary Table S10).

## DISCUSSION

Seasonal clock changes produced detectable disruptions in sleep, but the clearest evidence emerged in how sleep changed across nights rather than in sleep duration on a single post-transition night. Following the spring transition, sleep duration changed more from one night to the next in Mountain states than in Arizona, even though sleep duration itself on day +1 showed little evidence of a corresponding DST-related reduction. Sleep timing also shifted around both seasonal transitions. Among Mountain-state observations with a substantial post-transition departure from usual sleep duration, most returned to their baseline range within two nights, and greater baseline sleep variability predicted a greater likelihood of remaining outside that range at later checkpoints. Critically, however, a nearly identical two-night return pattern appeared in Arizona on the same calendar dates. Thus, although the spring clock change produced an acute disruption in sleep, the subsequent return-to-baseline trajectory cannot be interpreted as the time course of recovery from DST itself.

Aim 1 used two distinct outcomes. The first describes sleep duration on spring day +1 itself. The second describes how much that duration differed from the preceding observed night. On spring day +1, the Mountain-versus-Arizona difference in sleep duration was similar to that on day -1. The night-to-night change was 14.6 minutes larger in Mountain states relative to Arizona. The targeted analysis requiring an observed Sunday night immediately before Monday produced a similar 18.2-minute estimate. Because the same Sunday-to-Monday calendar transition occurred in Arizona, this result is unlikely to be explained by ordinary variation across the week alone. Wearable studies commonly show longer weekend sleep and shifts in sleep timing across the week.^9,10^ Our finding extends prior reports of acute spring sleep changes by showing that the clearest signal may appear in the change between nights even when sleep duration on the focal night changes little.^2,6–8^

Sleep timing provided a complementary Aim 1 signal. In spring, onset and midpoint were later in Mountain states relative to Arizona. In fall, onset, midpoint, and wake time shifted earlier by about 27-28 minutes, roughly half of the one-hour civil-time change. These partial shifts are consistent with an abrupt change in social time occurring against an unchanged light-dark cycle.^1^ Heacock et al.^8^ reported related changes in sleep timing around U.S. clock transitions, and Jeong et al.^17^ found a redistribution of physical activity timing using the same Arizona-versus-Mountain contrast. Across the broader literature, the clearest sleep effects are generally concentrated near the transition, particularly in spring, while the reported duration varies with the sleep measure and study design.^2,5^

A unique feature of our study is that we described return among Mountain-state windows that met the preregistered >30-minute criterion. The median return to the pre-transition ±30-minute window was two nights, and about one sixth of qualifying windows remained outside the window at day +7. Taken alone, this pattern suggests that large post-transition departures are usually short-lived. The post hoc Arizona analysis provides the critical context for that interpretation. Applying the same selection-and-return rule on matched Arizona dates produced almost the same qualifying rates, the same two-night median return, and an adjusted return rate close to 1.0 relative to Mountain states. The same-weekday analysis also left qualifying rates above 91%. Once a window is selected because it contains an extreme value relative to a recent baseline, subsequent observations are expected to move closer to that baseline through ordinary within-person variation and regression to the mean.^14^ The two-night median therefore describes the behavior of this threshold-defined return measure; it cannot be used to estimate the duration of circadian adaptation to daylight saving time.

Aim 3 adds an individual-difference perspective to that return measure. Greater pre-transition sleep-duration variability was consistently associated with remaining outside the baseline ±30-minute window at days +7 and +14. Short baseline sleep was associated with the day +7 outcome, while its day +14 confidence interval included 1. These results fit prior work treating sleep regularity as a dimension of sleep health that is distinct from mean duration.^11–13^ In this study, baseline variability appears to identify people whose sleep durations are more likely to remain far from their recent mean after a large departure. Direct circadian phase measures would be needed to determine whether the same characteristic predicts physiologic adaptation to a clock shift.

Our study contains several noteworthy strengths. The design uses repeated transitions and a neighboring no-clock-change comparison observed on the same calendar dates. Participant-transition fixed effects control stable characteristics within each transition window, and exact-date fixed effects absorb shocks shared across regions. The primary sleep processing followed a published All of Us algorithm, while targeted analyses tested weekday structure and an alternative Fitbit summary. Preregistration separated the primary questions from the post hoc specificity checks, allowing the Arizona analysis to be interpreted as a test of the return measure rather than as a second preregistered recovery hypothesis.

This work should also be understood in light of several limitations. All of Us Fitbit contributors are a selective, unweighted subset of program participants, limiting population representativeness. State of residence was a fixed participant-level field, so moves, travel, and transition-specific time zones were unavailable; available geography also could not isolate Navajo Nation residents in Arizona who observe daylight saving time. Some fall event-study outcomes showed differential pre-transition patterns. Fitbit sleep measures are device-derived and depend on episode-construction rules;^23,24^ the estimated magnitude of the spring night-to-night effect also differed across the two Fitbit sleep-processing approaches. The primary night-to-night measure can span a missing calendar night, although the consecutive Sunday-to-Monday analysis yielded a similar spring estimate. Missing nights may also delay the first observed return. The 30-minute threshold is operational and has no established clinical meaning. The return measure shares its baseline with the departure definition, creating regression-to-the-mean pressure and mathematical coupling; the Arizona analysis and unconditioned event study demonstrate how strongly this can shape interpretation. The saved default isMainSleep nightly data were insufficient to reconstruct the ordered Aim 2 return measure, so that pipeline sensitivity was limited to Aim 1.

## Conclusion

Seasonal clock changes produced a measurable acute shift in sleep timing and, in the spring, affected how much sleep duration changed from one observed night to the next. The study also shows that a visually plausible return-to-baseline curve can arise when the same threshold-based algorithm is applied on dates without a local clock change. For wearable studies of short-term recovery, the exposure effect and the statistical behavior of the return measure need to be evaluated separately. Comparison periods or populations that do not receive the exposure are therefore central to interpreting individual recovery trajectories.

## Data Availability

The data used in this study were obtained from the National Institutes of Health All of Us Research Program (Controlled Tier, Curated Data Repository version 8). Individual-level data are available to approved researchers through the secure All of Us Researcher Workbench, subject to applicable registration and data use requirements (https://www.researchallofus.org/data-tools/workbench/). Participant-level data cannot be publicly shared. Only aggregate results meeting All of Us dissemination requirements are reported in the manuscript and supplementary materials.

## Declarations

## Preregistration

The sleep-focused analysis plan was preregistered on the Open Science Framework (OSF): https://osf.io/52ucq/registrations.

## Transparency and openness

Preregistration deviations and post hoc additions, including the Arizona specificity analysis, are documented in Supplementary Table S1; data access and export restrictions are described below. The analytic code used in this study will be made available upon publication.

## Data availability

Row-level All of Us data are available to approved researchers through the secure Researcher Workbench. Only aggregate outputs meeting All of Us dissemination requirements are exportable.

## Declaration of generative AI use

During manuscript preparation, the authors used OpenAI ChatGPT for language editing and code troubleshooting. The authors made all analytic decisions, executed the analyses inside the All of Us Researcher Workbench, verified every reported result, and take responsibility for the final manuscript. Row-level All of Us data remained inside the secure Researcher Workbench.

## Author contributions (Contributor Roles Taxonomy [CRediT])

Conceptualization: Shuai Yang (SY), David A. Sbarra (DAS), Yann C. Klimentidis (YCK). Methodology: SY, DAS, YCK. Data curation: SY. Software: SY. Formal analysis: SY. Visualization: SY. Writing - original draft: SY. Writing - review and editing: SY, DAS, YCK. Supervision: DAS, YCK. SY had full access to the study data and is the guarantor.

## Ethics and dissemination

All of Us participants provided informed consent for electronic health record linkage and survey collection. The All of Us Research Program is reviewed and approved by the All of Us Institutional Review Board (AoU IRB Protocol Number: 2016-05). The present secondary analysis used de-identified Controlled Tier v8 data through the secure All of Us Researcher Workbench under a data use agreement, with aggregate reporting only.

## Acknowledgements

The authors gratefully acknowledge the All of Us Research Program participants, whose contributions made this work possible, and the All of Us Research Program staff and partners for maintaining the Researcher Workbench and data resources.

