## Supplementary Material for "Daylight Saving Time, Acute Sleep Disruption, and Return-to-Baseline Patterns in All of Us Fitbit Data"

**Supplementary Methods**

**Data extraction and daylight saving time calendar**

The analysis was conducted in the All of Us Researcher Workbench using Curated Data Repository version 8 (CDR v8) Controlled Tier data. Daylight saving time (DST) dates followed the post-2007 U.S. schedule. Eligible transitions began in spring 2017 and ended in spring 2023. A transition entered the analysis when its 21-day post-event processing window fell before the October 1, 2023 CDR cutoff. The preregistration-to-analysis crosswalk is provided in Table S1.

State of residence was obtained from the participant-level cb_search_person.state_of_residence field and linked to each participant. Arizona was coded as the no-DST comparison. Colorado, New Mexico, and Utah formed the DST-observing Mountain group. The recorded state field was held fixed across a participant's observed windows because transition-specific residence dates were unavailable.

**Typical sleep period (TSP) preprocessing**

We applied the published TSP algorithm to Fitbit sleep_level records for participants in the regional cohort. The query returned the stage-level fields required to reconstruct nightly sleep. TSP uses a participant's habitual primary sleep period to assign fragmented records to a nightly episode. The source panel began 120 days before the first eligible transition and ended 21 days after the last transition. This interval supplied available look-back history for TSP processing and did not impose a 120-night eligibility requirement. Participant-transition eligibility was determined from the nightly output.^1^

Sleep-level records were processed in participant chunks using the typical.sleep sequence as_sleep_logs(), as_typical_sleep(), and get_sleep_metrics(). The standardized nightly output included total sleep time, time in bed, sleep efficiency, sleep onset, sleep offset, and midpoint when available. BigQuery DATETIME values were parsed with an Etc/UTC label to retain the supplied clock values during processing.

Sleep efficiency values supplied as percentages were converted to proportions before validation. A valid night required 2-15 hours of sleep and efficiency between 50% and 100%. The default-pipeline sensitivity used sleep_daily_summary rows designated as isMainSleep.

**Participant-transition construction and baseline**

Nightly records were linked to the corresponding spring or fall transition and retained from event day -21 through +21. Each participant-transition received a unique identifier. Primary models used days -14 to -1 and +1 to +14; day 0 was excluded. The primary completeness rule required at least seven valid nights on each side of the transition. Baseline sleep duration was the mean across valid nights from day -14 through day -1, baseline efficiency used the same window, and sleep-duration standard deviation quantified baseline variability. At least four valid pre-transition nights were required to calculate a baseline before the 7-pre/7-post primary eligibility rule was applied.

Acute departure metrics used days +1 to +3. Short baseline sleep was defined as mean sleep duration <6 hours. Age at each transition was calculated from transition year and year of birth. The same participant could contribute multiple transition windows.

**Event-study outcomes and models**

Absolute deviation was the absolute difference between nightly sleep duration and the participant-transition mean baseline. Signed deviation subtracted the baseline from nightly duration. The primary adjacent-observation outcome was abs(sleep_minutes - lag(sleep_minutes)) after chronological ordering within participant-transition; lag() therefore referred to the preceding observed nightly record. This outcome was calculated before the event-study filter removed day 0 as a standalone observation, so an observed day 0 night could serve as the preceding night for the day +1 value. Sleep-efficiency outcomes used corresponding baseline operations.

For each outcome and season, Aim 1 used an event-study difference-in-differences specification estimated with fixest::feols(): outcome ~ i(event_day, observes_dst, ref = -1) | participant_transition_id + date. Standard errors were clustered by person_id. Each event-day coefficient compares the Mountain-versus-Arizona difference with the same regional difference on day -1. Joint pre-transition tests covered days -14 through -2. The Wald statistic was b'V^-1b and was evaluated against a chi-square distribution with 13 degrees of freedom.^2,3^ Complete joint tests are reported in Table S2. The targeted consecutive Sunday-to-Monday sensitivity required both nights in each pair to be valid and exactly one calendar day apart; it compared the Mountain-versus-Arizona contrast on post-transition Monday with the corresponding contrast on ordinary pre-transition Mondays at event days -13 and -6 (Table S3).

Timing analyses used TSP clock-time fields. Clock time was converted to minutes after midnight, and the pre-transition reference was calculated with a circular mean. Nightly change used the shortest signed circular difference from that reference, bounded at +/-720 minutes. At least four pre-transition timing observations were required. The event-study specification matched the primary Aim 1 models.

**Recovery analyses**

The primary return cohort included Mountain participant-transitions that met the 7-pre/7-post rule and crossed the 30-minute sleep-duration departure threshold during days +1 to +3. The first qualifying departure was the earliest valid night with an absolute departure >30 minutes from the participant-transition baseline. Follow-up began after that night. Return was the first subsequent valid night, through transition day +14, within 30 minutes of baseline. Time to return was the number of nights from first qualifying departure to subsequent return; windows without an observed return were censored at day +14. Prespecified unrecovered checkpoints remained indexed to transition day +3, +7, and +14. Alternative qualifying definitions used the same ordering and are reported in Table S4.

The Cox model was Surv(followup_time_nights, recovered_by_14) ~ scale(first_disturbance_magnitude_min) + transition + scale(baseline_sleep_minutes) + baseline_short_sleep_6h + cluster(person_id). First-departure magnitude was the absolute departure at the start of follow-up. Post hoc diagnostics summarize the timing and direction of the first qualifying departure (Table S5). The calendar-matched Arizona specificity analysis applied the identical qualifying and subsequent-return algorithm; results are reported in Table S6 and Figure S1. A same-weekday baseline sensitivity compared post-transition Monday, Tuesday, and Wednesday with the corresponding weekdays in the two pre-transition weeks and retained windows with all three comparisons available (Table S7). Figure S2 shows the unconditioned Mountain-versus-Arizona event study for absolute sleep-duration deviation without threshold selection.

**Baseline-predictor models**

The parsimonious Aim 3 model was unrecovered status ~ baseline short sleep + standardized baseline sleep-duration SD + standardized age + transition type + state. Colorado served as the state reference. Models were fitted separately for transition day +7 and day +14 using participant-clustered robust Wald inference. Full coefficients are reported in Table S8.

**Sensitivity analyses**

The typical sleep period versus default isMainSleep Aim 1 sensitivity is reported in Table S9, and the analysis excluding 2020 and 2021 transitions is reported in Table S10.

Analyses were run in R 4.5.0. Event-study models used the *fixest* package (Bergé et al., 2026),^4^ Cox models used the *survival* package,^5^ and nightly preprocessing used *typical.sleep*.^1^ Full package versions were recorded with sessionInfo() and retained in the analysis logs.

**Table S1. Preregistration-to-analysis crosswalk.**

| **Preregistered element** | **Planned specification** | **Implemented analysis** | **Status/clarification** |
| --- | --- | --- | --- |
| Aim 1: acute sleep disturbance | Arizona vs CO/NM/UT; spring and fall separately; sleep duration and regularity prioritized | Stacked event-study difference-in-differences with participant-transition and date fixed effects | Completed. Duration and adjacent-observation change reported as primary; efficiency and timing reported as secondary. Consecutive Sunday-to-Monday check added post hoc. |
| Aim 1: timing | Sleep midpoint when available; onset/offset as related outcomes | Circular clock-time event-study | Completed as secondary analyses after TSP timing fields were verified. |
| Aim 2: sleep recovery | Acute disturbance >30 min during days +1 to +3; first return within 30 min; days +3/+7/+14 | First qualifying >30-min departure during days +1 to +3; follow-up begins afterward; descriptive return, Cox, and binary delayed-return models. | First-departure ordering implemented. Arizona specificity check, departure-direction, and same-weekday checks added post hoc. |
| Aim 3: baseline vulnerability | Short sleep, sleep variability, age, midpoint, westward position; tighter regression if needed | Parsimonious logistic models for day +7/+14 | Short sleep, variability, and age included. Midpoint and westward position omitted because planned predictor definitions were unavailable. Transition and state included as design covariates. |
| Aim 3: exploratory discrimination | Discovery-oriented model with a held-out participant split | Not rerun | Exploratory analysis not rerun and not used in the manuscript conclusions. |
| Social determinants of health (SDoH) / social-material vulnerability (SMV) | Use only when measured before transition | Temporally ordered vulnerability analyses | Not included in this manuscript. |
| Navajo Nation sensitivity | Exclude or flag likely Navajo Nation 3-digit ZIP Code areas | Geographic sensitivity | Not run because ZIP3/county fields were unavailable. |
| Electronic health record (EHR) encounter check | Exploratory if feasible | Pre/post encounter-count analysis | Not included in this manuscript. |
| National extension | Secondary geographic contrast | Arizona/Hawaii vs DST-observing states | Not run; regional contrast retained as primary. |

**Table S2. Joint pre-transition tests for event-study models.**

| **Transition** | **Outcome** | **Days tested** | **df** | **Wald χ²** | **p** |
| --- | --- | --- | --- | --- | --- |
| Fall | Absolute deviation from baseline | -14, -13, -12, -11, -10, -9, -8, -7, -6, -5, -4, -3, -2 | 13 | 25.41 | 0.020 |
| Fall | Absolute change from preceding observed night | -14, -13, -12, -11, -10, -9, -8, -7, -6, -5, -4, -3, -2 | 13 | 11.26 | 0.589 |
| Fall | Sleep efficiency | -14, -13, -12, -11, -10, -9, -8, -7, -6, -5, -4, -3, -2 | 13 | 16.97 | 0.200 |
| Fall | Sleep duration | -14, -13, -12, -11, -10, -9, -8, -7, -6, -5, -4, -3, -2 | 13 | 22.64 | 0.046 |
| Spring | Absolute deviation from baseline | -14, -13, -12, -11, -10, -9, -8, -7, -6, -5, -4, -3, -2 | 13 | 14.88 | 0.315 |
| Spring | Absolute change from preceding observed night | -14, -13, -12, -11, -10, -9, -8, -7, -6, -5, -4, -3, -2 | 13 | 10.48 | 0.654 |
| Spring | Sleep efficiency | -14, -13, -12, -11, -10, -9, -8, -7, -6, -5, -4, -3, -2 | 13 | 9.38 | 0.744 |
| Spring | Sleep duration | -14, -13, -12, -11, -10, -9, -8, -7, -6, -5, -4, -3, -2 | 13 | 20.46 | 0.084 |

Note. Day -1 was the reference.

**Table S3. Targeted consecutive Sunday-to-Monday sensitivity for absolute night-to-night change.**

| **Season** | **Estimate, min** | **95% CI** | **p** | **Observations** | **Participants** | **PT** |
| --- | --- | --- | --- | --- | --- | --- |
| Spring | 18.2 | 11.2 to 25.3 | <.001 | 8,763 | 1,369 | 3,530 |
| Fall | -1.1 | -8.0 to 5.9 | .757 | 7,315 | 1,240 | 2,973 |

Note. The estimate is the Mountain-versus-Arizona excess on post-transition Monday relative to ordinary pre-transition Sunday-to-Monday pairs. Both nights in each pair were valid and exactly one calendar day apart. Models included participant-transition and exact-date fixed effects, with standard errors clustered by participant. CI = confidence interval; PT = participant-transitions.

**Table S4. Return to baseline under alternative qualifying-departure definitions.**

| Definition | Season | N | PT | Median nights | Unrecovered +7 | Unrecovered +14 |
| --- | --- | --- | --- | --- | --- | --- |
| Any night >30 min (primary) | Spring | 712 | 1476 | 2 | 17.1% | 3.3% |
| Any night >30 min (primary) | Fall | 617 | 1213 | 2 | 16.2% | 3.3% |
| Any night >45 min | Spring | 662 | 1273 | 2 | 19.1% | 3.8% |
| Any night >45 min | Fall | 573 | 1049 | 2 | 18.6% | 4.0% |
| Mean acute deviation >30 min | Spring | 654 | 1228 | 3 | 19.1% | 3.9% |
| Mean acute deviation >30 min | Fall | 565 | 1013 | 2 | 18.0% | 3.9% |
| At least two nights >30 min | Spring | 580 | 996 | 3 | 20.2% | 4.2% |
| At least two nights >30 min | Fall | 490 | 810 | 3 | 18.8% | 4.2% |
| Any night >0.5 baseline SD | Spring | 709 | 1479 | 2 | 16.5% | 3.2% |
| Any night >0.5 baseline SD | Fall | 607 | 1210 | 2 | 16.0% | 3.3% |

Note. All definitions enforce temporal ordering: the return event must occur after the qualifying departure. Unrecovered checkpoints are indexed to the transition date; median time is measured from the first qualifying departure. N = participants; PT = participant-transitions.

**Table S5. Direction and timing of the first qualifying >30-minute departure.**

| Region | Season | Qualifying PT | First on day +1 | First on day +2 | First on day +3 | Shorter first | Longer first |
| --- | --- | --- | --- | --- | --- | --- | --- |
| Mountain DST | Spring | 1,476 | 93.8% | 5.1% | 1.1% | 51.7% | 48.3% |
| Mountain DST | Fall | 1,213 | 94.5% | 4.9% | 0.7% | 54.1% | 45.8% |
| Arizona no-DST | Spring | 1,726 | 91.0% | 7.6% | 1.4% | 50.8% | 49.2% |
| Arizona no-DST | Fall | 1,520 | 91.9% | 6.7% | 1.4% | 52.8% | 47.2% |

Note. The qualifying departure is the earliest valid night on days +1 to +3 with an absolute sleep-duration departure >30 minutes from the participant-transition mean baseline. Direction is based on the signed difference from baseline. PT = participant-transitions; DST = daylight saving time.

**Table S6. Post hoc Arizona calendar-matched specificity analysis using the same first-departure algorithm.**

| Region | Season | Qualifying / eligible PT | Qualifying rate | Median nights | Unrecovered +7 | Unrecovered +14 |
| --- | --- | --- | --- | --- | --- | --- |
| Mountain DST | Spring | 1476 / 1660 | 88.9% | 2 | 17.1% | 3.3% |
| Mountain DST | Fall | 1213 / 1363 | 89.0% | 2 | 16.2% | 3.3% |
| Arizona no-DST | Spring | 1726 / 1952 | 88.4% | 2 | 17.6% | 4.2% |
| Arizona no-DST | Fall | 1520 / 1683 | 90.3% | 2 | 17.4% | 3.9% |

Note. The curves were nearly identical. In a participant-clustered Cox model among qualifying windows, Mountain daylight saving time versus Arizona no-DST HR = 1.03 (95% CI [0.98, 1.09], p = .247), adjusted for transition, age, baseline sleep duration, and baseline sleep-duration variability. HR = hazard ratio; CI = confidence interval. PT = participant-transitions.

**Table S7. Same-weekday baseline sensitivity for the >30-minute qualifying departure.**

| Region | Season | Eligible PT | Participants | Qualifying PT | Qualifying rate | Shorter first | Longer first |
| --- | --- | --- | --- | --- | --- | --- | --- |
| Mountain DST | Spring | 1,069 | 539 | 989 | 92.5% | 51.4% | 48.6% |
| Mountain DST | Fall | 857 | 461 | 787 | 91.8% | 52.2% | 47.6% |
| Arizona no-DST | Spring | 1,090 | 441 | 1,007 | 92.4% | 47.6% | 52.3% |
| Arizona no-DST | Fall | 932 | 424 | 872 | 93.6% | 48.7% | 51.3% |

Note. Each post-transition day was compared with the mean of available valid observations from the same weekday in the two pre-transition weeks: day +1 Monday with days -13 and -6, day +2 Tuesday with days -12 and -5, and day +3 Wednesday with days -11 and -4. The table is restricted to participant-transitions with all three post-transition comparisons available. PT = participant-transitions; DST = daylight saving time.

**Table S8. Full baseline-predictor logistic models for day +7 and day +14 unrecovered status.**

| **Outcome** | **Predictor** | **OR** | **95% CI low** | **95% CI high** | **p** |
| --- | --- | --- | --- | --- | --- |
| Day +7 | Baseline short sleep <6 h | 1.56 | 1.13 | 2.15 | .007 |
| Day +7 | Baseline sleep-duration SD, per SD | 1.77 | 1.60 | 1.95 | <.001 |
| Day +7 | Age, per SD | 0.87 | 0.77 | 0.97 | .016 |
| Day +7 | Spring vs fall | 1.14 | 0.92 | 1.41 | .233 |
| Day +7 | New Mexico vs Colorado | 0.87 | 0.65 | 1.17 | .362 |
| Day +7 | Utah vs Colorado | 0.85 | 0.61 | 1.19 | .340 |
| Day +14 | Baseline short sleep <6 h | 1.78 | 0.99 | 3.18 | .053 |
| Day +14 | Baseline sleep-duration SD, per SD | 1.94 | 1.66 | 2.26 | <.001 |
| Day +14 | Age, per SD | 0.86 | 0.67 | 1.10 | .232 |
| Day +14 | Spring vs fall | 1.07 | 0.70 | 1.64 | .761 |
| Day +14 | New Mexico vs Colorado | 1.16 | 0.63 | 2.16 | .636 |
| Day +14 | Utah vs Colorado | 0.78 | 0.41 | 1.47 | .442 |

Note. Odds ratios and 95% confidence intervals use participant-clustered robust Wald inference. Colorado and fall are the reference categories. Models include 2,689 participant-transitions from 799 participants.

**Table S9. Selected TSP versus default isMainSleep Aim 1 sensitivity results.**

| Pipeline | Quantity | Estimate | p |
| --- | --- | --- | --- |
| TSP | Primary completeness sample | 1,522 participants; 6,658 PT |  |
| Default isMainSleep | Primary completeness sample | 2,021 participants; 8,666 PT |  |
| TSP | Spring day +1 sleep duration | -8.4 (-17.3 to 0.6) min | p = .066 |
| TSP | Spring day +1 change from preceding observed night | 14.6 (7.2 to 22.0) min | p < .001 |
| Default isMainSleep | Spring day +1 sleep duration | -4.4 (-12.4 to 3.6) min | p = .282 |
| Default isMainSleep | Spring day +1 change from preceding observed night | 7.0 (-0.3 to 14.2) min | p = .060 |

Note. Aim 1 estimates are shown for both nightly preprocessing pipelines. TSP = typical sleep period; PT = participant-transitions.

**Table S10. Sensitivity analysis excluding 2020 and 2021 transition years.**

| Analysis | Quantity | Result | p |
| --- | --- | --- | --- |
| Event-study | Sample | 1,410 participants; 4,375 PT |  |
| Recovery cohort | Sample | 749 participants; 1,848 PT |  |
| Event-study | Spring day +1 change from preceding observed night | 10.3 (1.7 to 18.9) min | p = .018 |
| Return to baseline | Fall recovery | median 2 nights; 15.3% unrecovered at +7; 3.5% at +14 |  |
| Return to baseline | Spring recovery | median 2 nights; 16.2% unrecovered at +7; 3.3% at +14 |  |

Note. Recovery uses the first-departure temporal ordering. The spring/fall unrecovered-at-+7 estimates were 16.2% and 15.3%, respectively, close to the primary analysis. PT = participant-transitions.

**Figure S1. Arizona no-DST versus Mountain daylight saving time cumulative return after the first qualifying >30-minute departure. The identical selection and subsequent-return algorithm was applied to calendar-matched windows in both regions. The near-overlapping curves and adjusted Mountain-versus-Arizona hazard ratio = 1.03 (95% confidence interval [0.98, 1.09], p = .247) indicate that the selected return pattern is not specific to daylight saving time.**

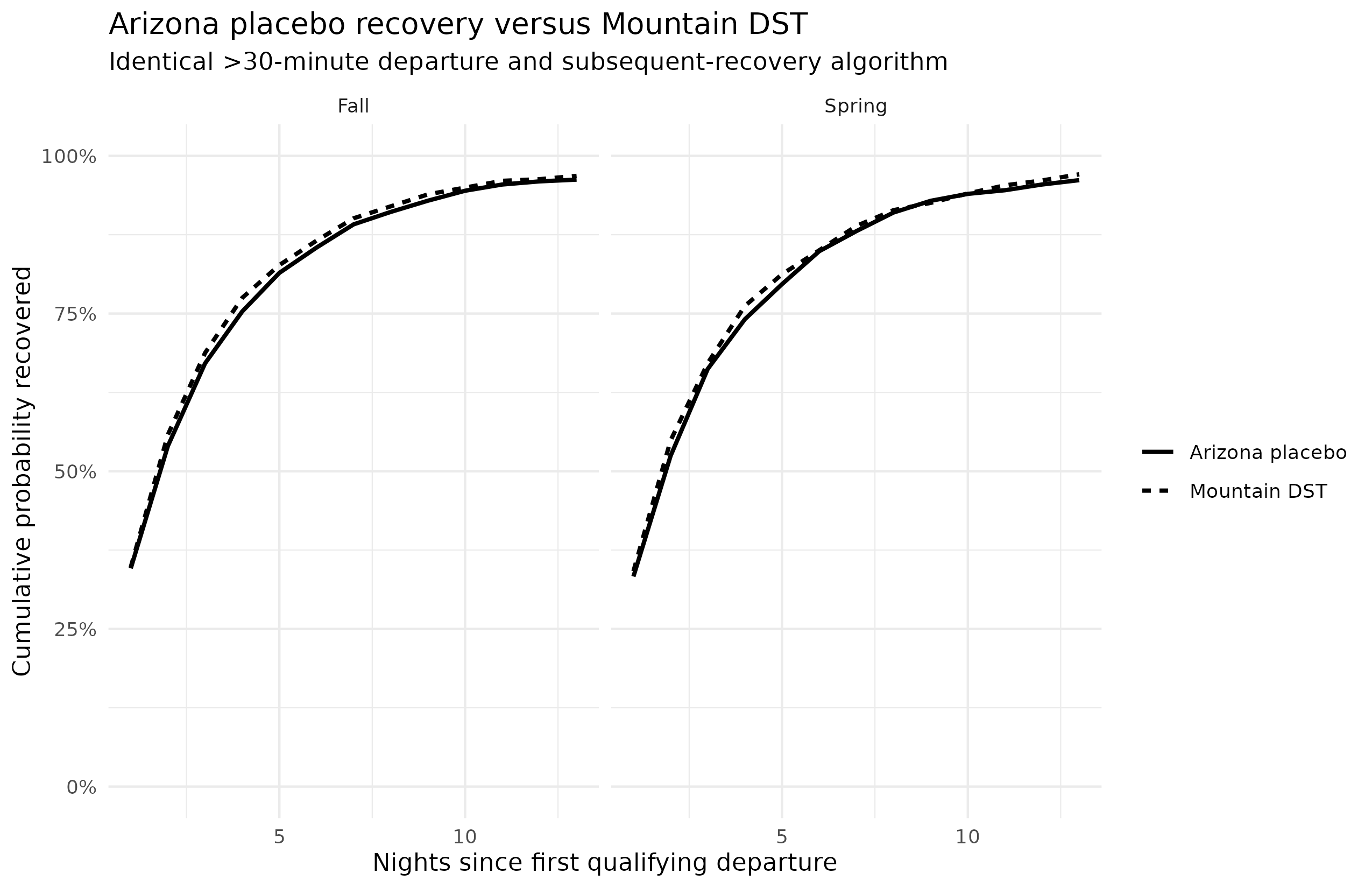

**Figure S2. Unconditioned Mountain-versus-Arizona event-study estimates for absolute sleep-duration deviation from participant-transition baseline.** No >30-minute departure selection was applied. Points show the Mountain-versus-Arizona contrast relative to day -1 and shaded bands are 95% confidence intervals; no clear post-transition excess absolute deviation was detected.

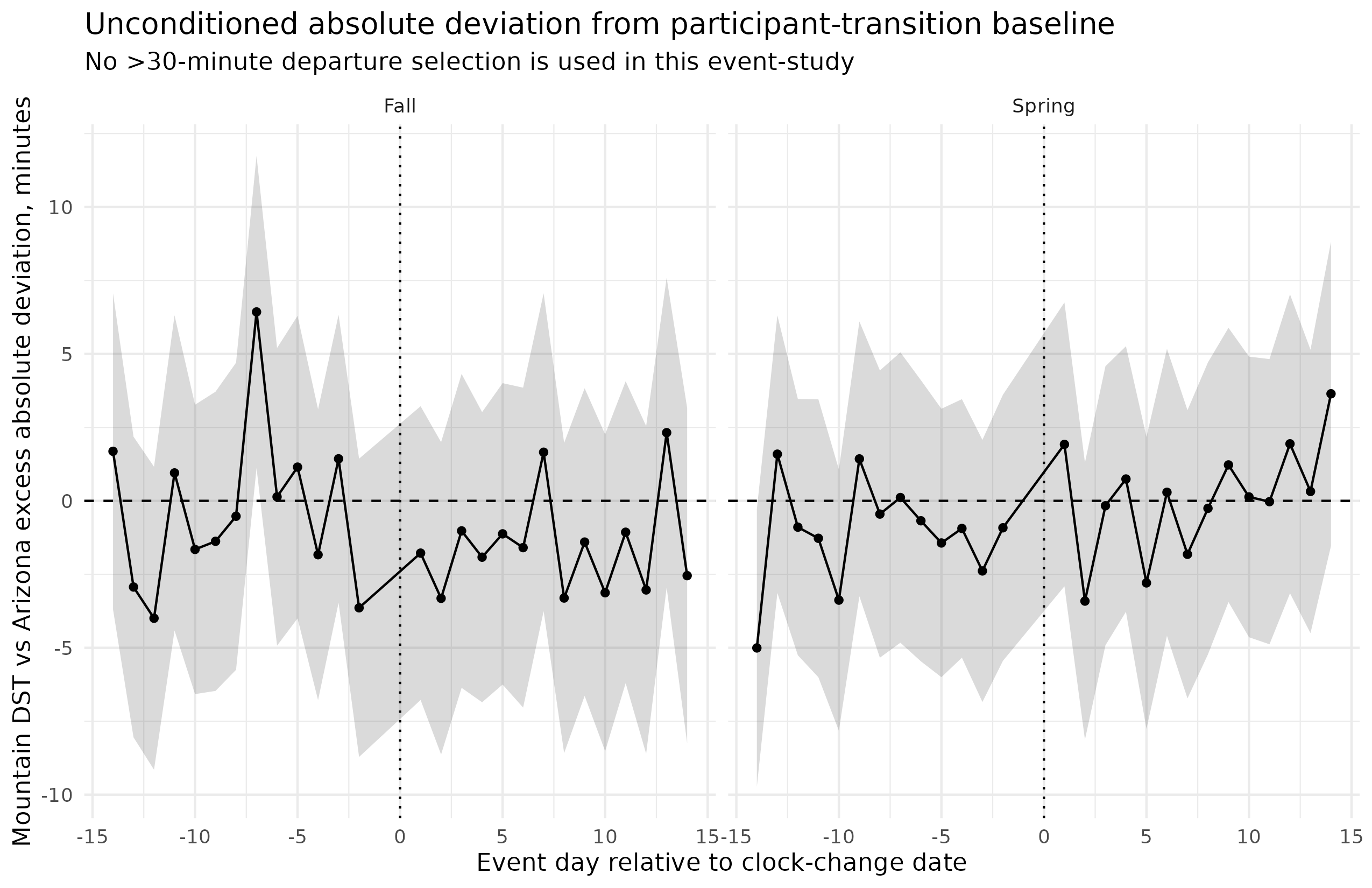
